# Antithrombotic Therapy in Atrial Fibrillation Patients with Prior Complex Percutaneous Coronary Intervention: A Secondary Analysis of the Randomized ADAPT AF-DES Trial

**DOI:** 10.64898/2026.02.26.26347227

**Authors:** Hong-Ki Jeon, Ho Sung Jeon, Kyounghoon Lee, Yun-Hyeong Cho, Cheol Ung Choi, Sang-Rok Lee, Hyung-Bok Park, Han Cheol Lee, Seunghwan Kim, Sang-Hyup Lee, Yong-Joon Lee, Seung-Jun Lee, Hee Tae Yu, Sung-Jin Hong, Chul-Min Ahn, Byeong-Keuk Kim, Young-Guk Ko, Donghoon Choi, Myeong-Ki Hong, Yangsoo Jang, Hui-Nam Pak, Jung-Sun Kim, Sung Gyun Ahn, ADAPT AF-DES investigators

## Abstract

**Background:** In patients with atrial fibrillation (AF) and stable coronary artery disease beyond 1 year after percutaneous coronary intervention (PCI), oral anticoagulant monotherapy is guideline-recommended; however, its efficacy and safety in patients with complex PCI remain uncertain.

**Methods:** We conducted a post-hoc analysis of the randomized ADAPT AF-DES trial comparing NOAC monotherapy versus NOAC plus clopidogrel in AF patients ≥12 months after second- or third-generation drug-eluting stent implantation. Complex PCI was defined by one of the following characteristics: ≥3 stents, ≥3 lesions, bifurcation with 2 stents, total stent length ≥60 mm, left main PCI, or chronic total occlusion PCI. Net adverse clinical events (NACE), ischemic composite outcomes, and bleeding composite outcomes were evaluated according to PCI complexity.

**Results:** Among 960 patients, 247 (25.7%) underwent complex PCI and 713 (74.3%) underwent noncomplex PCI. NOAC monotherapy was associated with a lower risk of NACE compared with combination therapy in both the complex PCI group (9.5% vs 21.5%; hazard ratio 0.42, 95% confidence interval 0.21–0.83; *P*=0.01) and the noncomplex PCI group (9.6% vs 15.7%; hazard ratio 0.59, 95% confidence interval 0.39–0.90; *P*=0.02), with no significant interaction. Ischemic outcomes did not differ significantly between treatment strategies regardless of PCI complexity, whereas bleeding outcomes were consistently lower with NOAC monotherapy in both complex and noncomplex PCI groups.

**Conclusions:** In this post hoc analysis of the randomized ADAPT AF-DES trial, the clinical benefits of NOAC monotherapy beyond 12 months after PCI—characterized by reduced bleeding without a significant increase in ischemic events—were consistent regardless of PCI complexity. While hypothesis-generating, these findings support a long-term antithrombotic strategy prioritizing bleeding reduction in patients with AF, irrespective of prior PCI complexity.

**Trial registration:** URL: http://www.clinicaltrials.gov; Unique identifier: NCT04250116.

**Clinical perspective:** *What is new?:*

- In a randomized population of patients with AF and prior drug-eluting stent implantation, the efficacy and safety of NOAC monotherapy versus NOAC plus clopidogrel were evaluated according to anatomic PCI complexity.
- Among patients with prior complex PCI, NOAC monotherapy was not associated with an increased risk of ischemic events and was associated with a substantial reduction in bleeding.

*What are the clinical implications?:*

- NOAC monotherapy beyond 1 year after PCI was supported in patients with AF, including those with prior complex PCI.
- Long-term antithrombotic decisions may place greater emphasis on bleeding risk than PCI complexity.
- The optimal duration of combination antithrombotic therapy after complex PCI in patients with AF remains to be determined.

## INTRODUCTION

Several randomized clinical trials and current practice guidelines indicate that, in patients with atrial fibrillation (AF) and stable coronary artery disease (CAD), oral anticoagulant monotherapy provides favorable outcomes compared with dual antithrombotic therapy by reducing bleeding risk without compromising ischemic protection.^1-4^ However, prior studies often included heterogeneous revascularization strategies such as coronary artery bypass grafting or bare-metal stents, and rarely focused on patients undergoing complex percutaneous coronary intervention (PCI).^1,2,5^ Consequently, evidence supporting guideline recommendations in AF patients treated with complex PCI remains limited.

Complex PCI—characterized by bifurcation stenting, chronic total occlusion intervention, left main PCI, or implantation of multiple or long stents— is associated with an increased risk of ischemic complications, including myocardial infarction and stent thrombosis.^6,7^ As a result, concerns regarding late thrombotic events may lead clinicians to hesitate in discontinuing antiplatelet therapy in patients with AF who have undergone complex PCI, despite the well-established bleeding risk associated with prolonged combination therapy.

The Appropriate Duration of Antiplatelet and Thrombotic Strategy after 12 Months in Patients with Atrial Fibrillation Treated with Drug-Eluting Stents (ADAPT AF-DES) trial demonstrated that non–vitamin K antagonist oral anticoagulant (NOAC) monotherapy reduced the composite risk of death, thromboembolic events, and major or clinically relevant nonmajor bleeding compared with NOAC plus clopidogrel among patients with AF and chronic CAD more than one year after drug-eluting stent implantation.^8^ In this post-hoc analysis of the ADAPT AF-DES trial, we investigated whether the clinical benefit of NOAC monotherapy is maintained in AF patients with prior complex PCI.

## METHODS

### Study design and population

The ADAPT AF-DES trial was a multicenter, randomized, open-label, noninferiority trial that enrolled 960 patients from 32 centers in South Korea. Eligible patients were aged ≥19 years, had a diagnosis of AF with a CHA₂DS₂-VASc score ≥2, and had undergone PCI with contemporary drug-eluting stents at least 1 year before enrollment. Key exclusion criteria included age >85 years; the need for anticoagulation for a mechanical prosthetic valve, moderate-to-severe mitral stenosis, or deep vein thrombosis; prior PCI with a first-generation drug-eluting stent; and a history of coagulopathy, recurrent bleeding, or a hemoglobin level <10 g/dL. Additional details of the study design and rationale have been published previously.^8^ In the present post-hoc analysis, the entire intention-to-treat population of the ADAPT AF-DES trial was evaluated to determine whether the treatment effects differed according to PCI complexity.

### Randomization and follow-up

Patients were randomly assigned in a 1:1 ratio to receive NOAC monotherapy or combination therapy (NOAC plus clopidogrel). Randomization was stratified by clinical presentation at PCI (stable CAD or acute coronary syndrome), NOAC type, HAS-BLED score (≥3 vs <3), and time from PCI to randomization (<24 vs ≥24 months).^9^ Patients in the monotherapy group received apixaban (5 mg twice daily) or rivaroxaban (20 mg once daily). Patients in the combination-therapy group received apixaban (5 mg twice daily) or rivaroxaban (15 mg once daily) plus clopidogrel (75 mg once daily). Dose reductions criteria have been previously described.^8^

Clinical follow-up was conducted at 6 and 12 months after randomization and included assessment of general health status, cardiovascular medication use, and the occurrence of trial end-point or adverse events.

### Definition of complex PCI and study endpoints

Complex PCI was defined according to the criteria used in the ADAPT AF-DES study, requiring the presence of at least one of the following complex features: implantation of ≥3 stents, treatment of ≥3 lesions, bifurcation PCI with two stents, total stent length ≥60 mm, left main lesion PCI, or chronic total occlusion PCI.^6-8,10^ PCI complexity was further quantified according to the number of complex features present in each patient: multiple complex PCI defined as ≥2 features.^6^

The primary endpoint was net adverse clinical events (NACE), defined as a composite of death from any cause, myocardial infarction, stent thrombosis, ischemic or hemorrhagic stroke, systemic embolism, and major bleeding or clinically relevant nonmajor bleeding at 12 months. Major bleeding or clinically relevant nonmajor bleeding were defined according to the criteria of the International Society on Thrombosis and Haemostasis.^11,12^

Secondary endpoints included ischemic outcomes (cardiovascular death, myocardial infarction, stent thrombosis, ischemic stroke, or systemic embolism) and bleeding outcomes (major or clinically relevant nonmajor bleeding). Individual components were also analyzed separately. Further definitions of the outcomes have been previously described.^8^

### Statistical analysis

Categorical variables are presented as numbers and percentages, and continuous variables are presented as means with standard deviations or medians with interquartile ranges, as appropriate. Comparisons of categorical variables between the two groups were performed using the chi-square test or Fisher’s exact test, as appropriate, and continuous variables were compared using the Student’s t-test or Mann–Whitney U test, depending on their distribution.

Time-to-event outcomes were evaluated from the date of randomization to the first event occurrence. Kaplan–Meier survival curves were generated, and event rates between treatment groups were compared using the log-rank test. Hazard ratios (HRs) with 95% confidence intervals (CIs) were estimated using unadjusted Cox proportional-hazards models. Additional subgroup analyses were conducted according to PCI complexity scores and individual complexity components. Multivariable Cox models adjusting for clinically relevant covariates (including age, sex, and HAS-BLED score) were applied in subgroup analyses to assess treatment effects across PCI complexity strata. Treatment-by-PCI complexity interactions were assessed by including interaction terms in the Cox models. All analyses were performed according to the intention-to-treat principle. All tests were two-sided, and *P* values < 0.05 were considered statistically significant. Statistical analyses were performed using R statistical software version 4.2.2 (R Foundation for Statistical Computing, Vienna, Austria).

## RESULTS

### Baseline characteristics

The study flow is presented in **Figure 1**. Of the 960 randomized patients enrolled in the ADAPT AF-DES trial, 247 (25.7%) underwent complex PCI and 713 (74.3%) underwent noncomplex PCI. The prevalence of the complex PCI components in the overall population is illustrated in **Figure 2**. Baseline characteristics were well balanced between the NOAC monotherapy and combination therapy groups within both PCI complexity strata (**Table 1**). Among patients with complex PCI, 126 were assigned to the NOAC monotherapy group and 121 to the combination therapy group, whereas among those with noncomplex PCI, 356 and 357 patients were assigned to each group, respectively.

**Figure 1.**
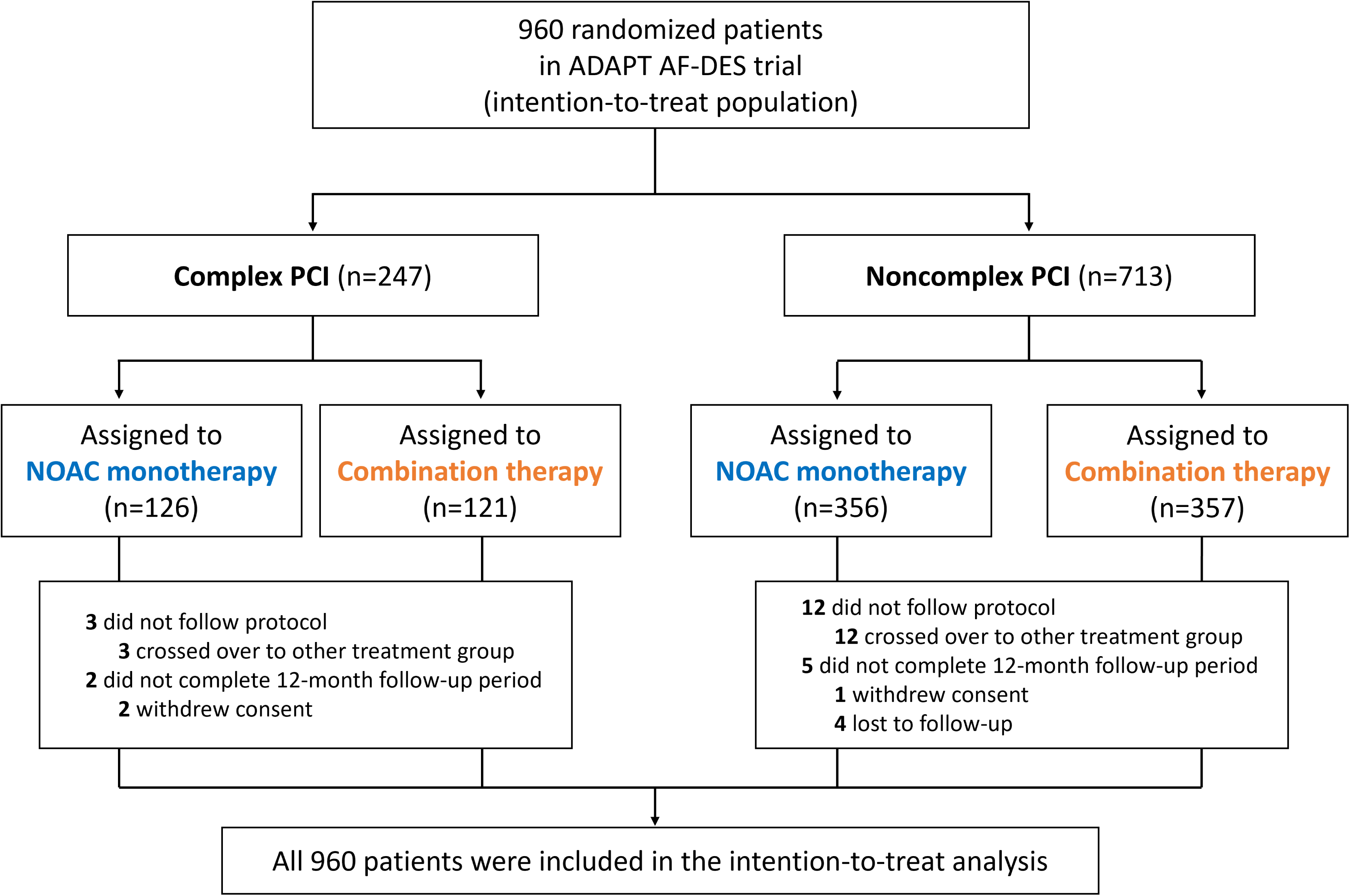
Study flowchart. A total of 960 patients were stratified by PCI complexity (complex vs noncomplex PCI). NOAC monotherapy was compared with combination therapy (NOAC plus clopidogrel) within each PCI complexity group. ADAPT AF-DES = Appropriate duration of antiplatelet and thrombotic strategy after 12 months in patients with atrial fibrillation treated with drug-eluting stents; NOAC = non-vitamin K antagonist oral anticoagulant; PCI = percutaneous coronary intervention.

**Figure 2.**
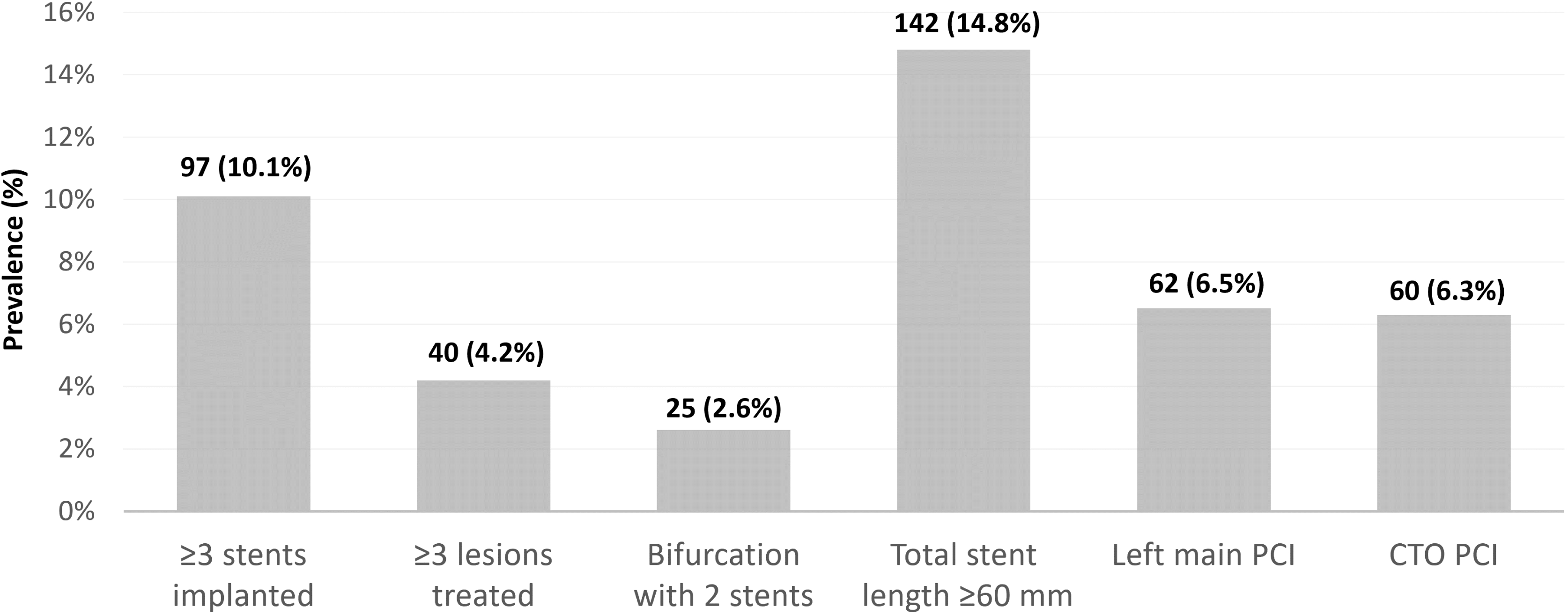
Prevalence of the complex PCI components in the overall population. Values are n (%). CTO = chronic total occlusion; PCI = percutaneous coronary intervention.

**Table 1.**
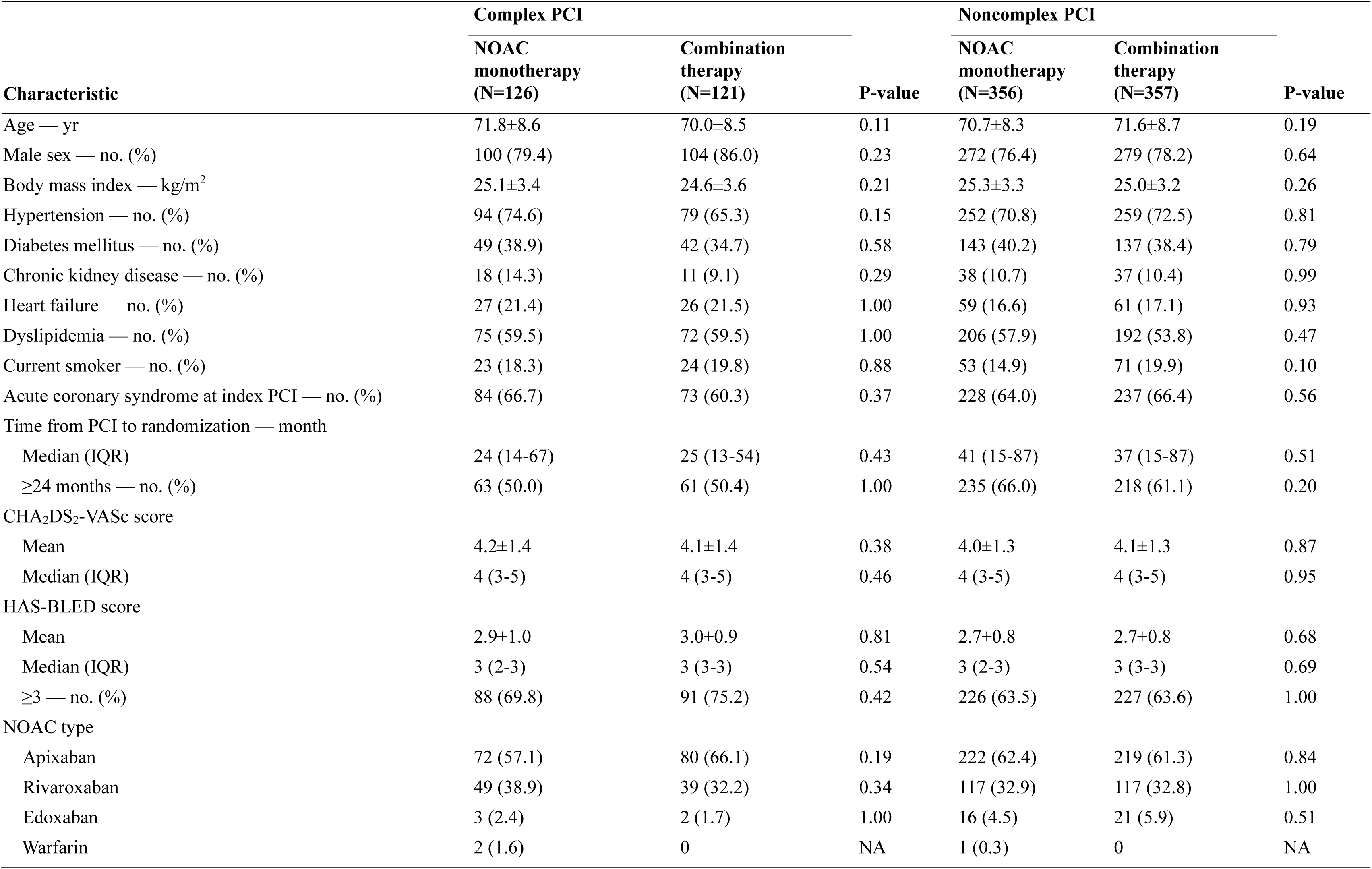
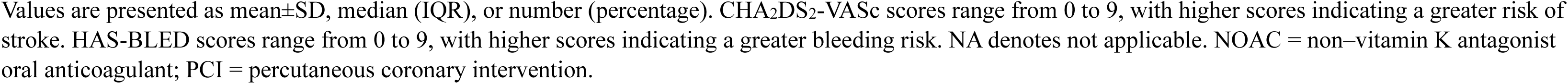
Baseline Characteristics.

| Characteristic | Complex PCI |  |  | Noncomplex PCI |  |  |
| --- | --- | --- | --- | --- | --- | --- |
|  | NOAC<br>monotherapy<br>(N=126) | Combination<br>therapy<br>(N=121) | P-value | NOAC<br>monotherapy<br>(N=356) | Combination<br>therapy<br>(N=357) | P-value |
| Age — yr | 71.8±8.6 | 70.0±8.5 | 0.11 | 70.7±8.3 | 71.6±8.7 | 0.19 |
| Male sex — no. (%) | 100 (79.4) | 104 (86.0) | 0.23 | 272 (76.4) | 279 (78.2) | 0.64 |
| Body mass index — kg/m<sup>2</sup> | 25.1±3.4 | 24.6±3.6 | 0.21 | 25.3±3.3 | 25.0±3.2 | 0.26 |
| Hypertension — no. (%) | 94 (74.6) | 79 (65.3) | 0.15 | 252 (70.8) | 259 (72.5) | 0.81 |
| Diabetes mellitus — no. (%) | 49 (38.9) | 42 (34.7) | 0.58 | 143 (40.2) | 137 (38.4) | 0.79 |
| Chronic kidney disease — no. (%) | 18 (14.3) | 11 (9.1) | 0.29 | 38 (10.7) | 37 (10.4) | 0.99 |
| Heart failure — no. (%) | 27 (21.4) | 26 (21.5) | 1.00 | 59 (16.6) | 61 (17.1) | 0.93 |
| Dyslipidemia — no. (%) | 75 (59.5) | 72 (59.5) | 1.00 | 206 (57.9) | 192 (53.8) | 0.47 |
| Current smoker — no. (%) | 23 (18.3) | 24 (19.8) | 0.88 | 53 (14.9) | 71 (19.9) | 0.10 |
| Acute coronary syndrome at index PCI — no. (%) | 84 (66.7) | 73 (60.3) | 0.37 | 228 (64.0) | 237 (66.4) | 0.56 |
| Time from PCI to randomization — month |  |  |  |  |  |  |
| Median (IQR) | 24 (14-67) | 25 (13-54) | 0.43 | 41 (15-87) | 37 (15-87) | 0.51 |
| ≥24 months — no. (%) | 63 (50.0) | 61 (50.4) | 1.00 | 235 (66.0) | 218 (61.1) | 0.20 |
| CHA<sub>2</sub>DS<sub>2</sub>-VASc score |  |  |  |  |  |  |
| Mean | 4.2±1.4 | 4.1±1.4 | 0.38 | 4.0±1.3 | 4.1±1.3 | 0.87 |
| Median (IQR) | 4 (3-5) | 4 (3-5) | 0.46 | 4 (3-5) | 4 (3-5) | 0.95 |
| HAS-BLED score |  |  |  |  |  |  |
| Mean | 2.9±1.0 | 3.0±0.9 | 0.81 | 2.7±0.8 | 2.7±0.8 | 0.68 |
| Median (IQR) | 3 (2-3) | 3 (3-3) | 0.54 | 3 (2-3) | 3 (3-3) | 0.69 |
| ≥3 — no. (%) | 88 (69.8) | 91 (75.2) | 0.42 | 226 (63.5) | 227 (63.6) | 1.00 |
| NOAC type |  |  |  |  |  |  |
| Apixaban | 72 (57.1) | 80 (66.1) | 0.19 | 222 (62.4) | 219 (61.3) | 0.84 |
| Rivaroxaban | 49 (38.9) | 39 (32.2) | 0.34 | 117 (32.9) | 117 (32.8) | 1.00 |
| Edoxaban | 3 (2.4) | 2 (1.7) | 1.00 | 16 (4.5) | 21 (5.9) | 0.51 |
| Warfarin | 2 (1.6) | 0 | NA | 1 (0.3) | 0 | NA |

Values are presented as mean±SD, median (IQR), or number (percentage). CHA<sub>2</sub>DS<sub>2</sub>-VASc scores range from 0 to 9, with higher scores indicating a greater risk of stroke. HAS-BLED scores range from 0 to 9, with higher scores indicating a greater bleeding risk. NA denotes not applicable. NOAC = non-vitamin K antagonist oral anticoagulant; PCI = percutaneous coronary intervention.

Comparisons between patients with complex and noncomplex PCI are summarized in **Supplementary Table S1**. Patients with complex PCI had higher bleeding risk, reflected by a higher HAS-BLED score (mean [SD], 2.9 [1.0] vs 2.7 [0.8]; *P*<0.001; median [IQR], 3 [2–3] vs 3 [2–3]; *P*<0.001) and a greater prevalence of high bleeding risk (HAS-BLED score ≥3: 72.5% vs 63.5%; *P*<0.001). The time from PCI to randomization was shorter in the complex PCI group than in the noncomplex PCI group (median [IQR], 24 months [14–60] vs 39 months [15–87]; *P*<0.001).

### Primary and secondary endpoints

Primary and secondary endpoints stratified by PCI complexity and antithrombotic strategy are summarized in **Table 2**, with Kaplan–Meier curves shown in **Figure 3**. Among patients with complex PCI, NOAC monotherapy was associated with a lower risk of NACE than combination therapy (9.5% vs. 21.5%; HR, 0.42; 95% CI, 0.21–0.83; *P*=0.01). A similar benefit was observed among those with noncomplex PCI (9.6% vs 15.7%; HR, 0.59; 95% CI, 0.39–0.90; *P*=0.02), without a significant interaction between PCI complexity and antithrombotic strategy (*P*_interaction_=0.40).

**Figure 3.**
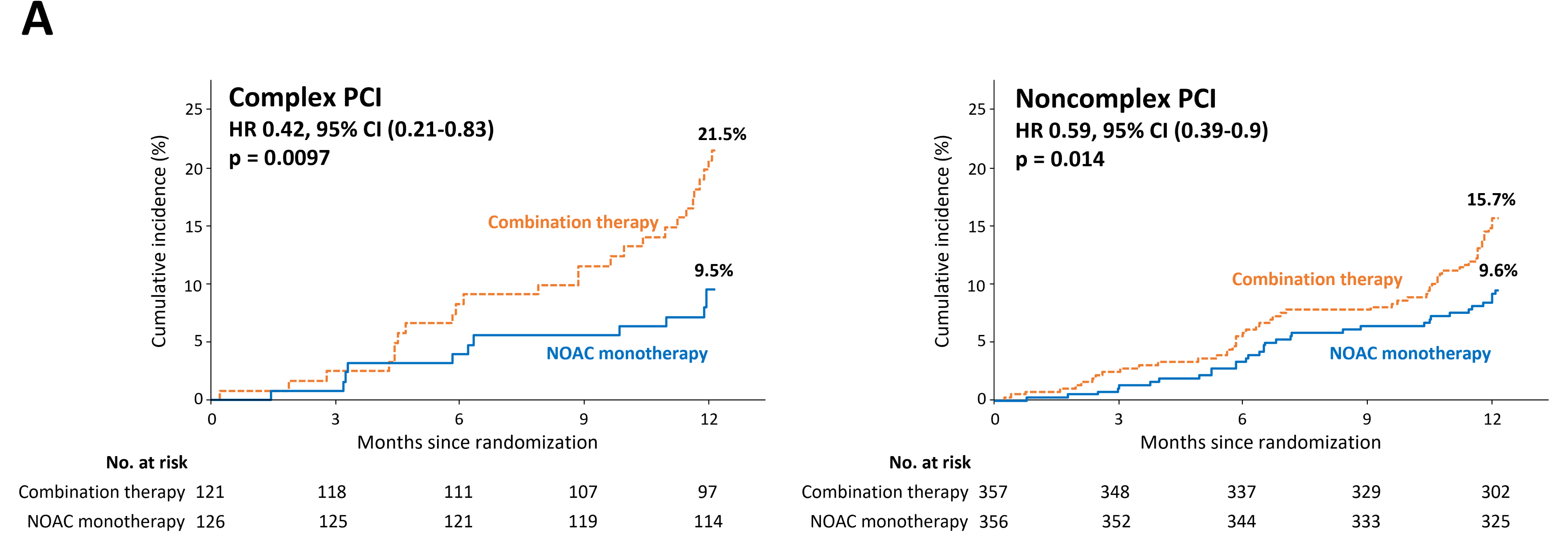
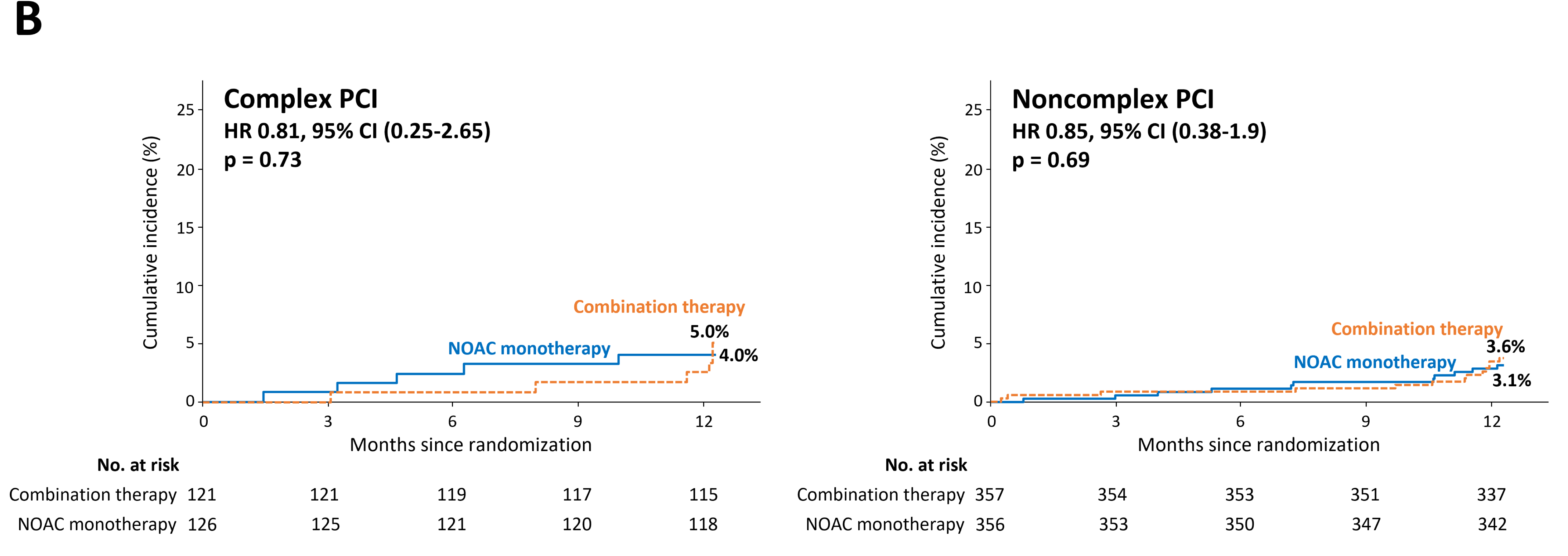
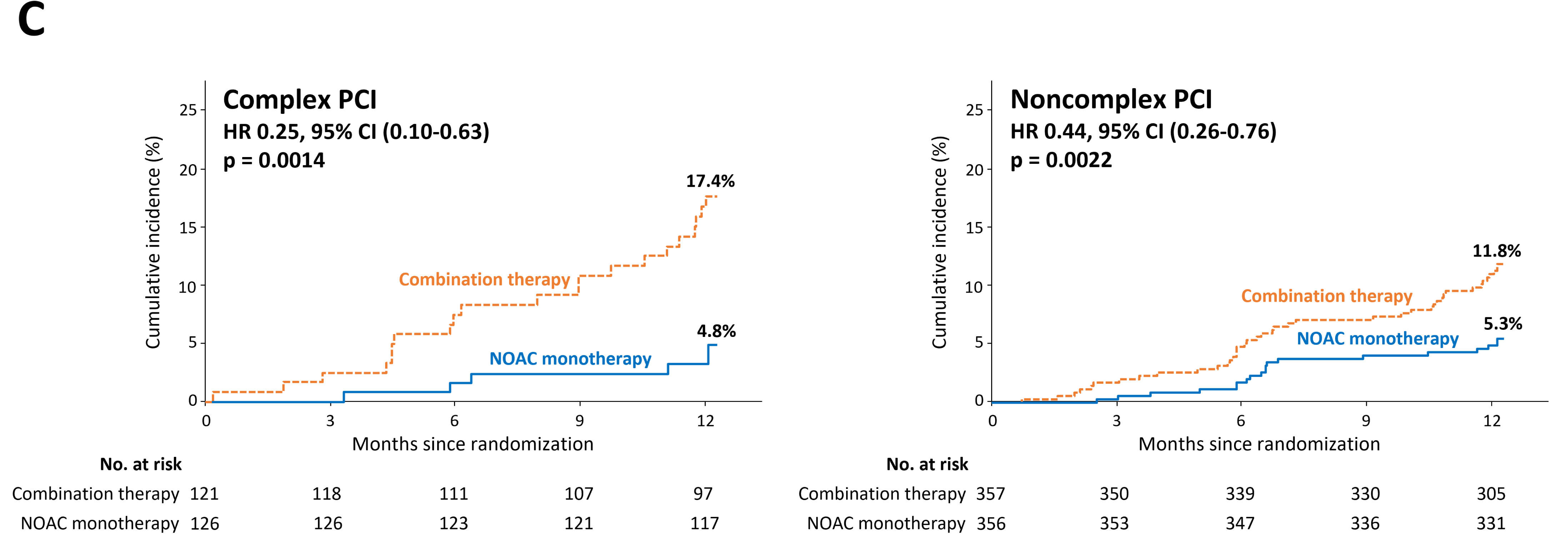
Kaplan–Meier curves stratified by PCI complexity. Time-to-event curves comparing NOAC monotherapy versus combination therapy (NOAC plus clopidogrel) for (A) the primary end point, net adverse clinical events (NACE; a composite of death from any cause, myocardial infarction, stent thrombosis, ischemic or hemorrhagic stroke, systemic embolism, and major or clinically relevant nonmajor bleeding), (B) ischemic composite outcomes (cardiovascular death, myocardial infarction, stent thrombosis, ischemic stroke, or systemic embolism), and (C) bleeding composite outcomes (major or clinically relevant nonmajor bleeding), stratified by complex and noncomplex PCI. HR = hazard ratio; CI = confidence interval. Other abbreviations as in **Figure 1 and 2**.

**Table 2.** Clinical outcomes.

|  | *No. (%)* |  |  |  | *No. (%)* |  |  |  |  |
| --- | --- | --- | --- | --- | --- | --- | --- | --- | --- |
|  | **Complex PCI** |  |  |  | **Noncomplex PCI** |  |  |  |  |
|  | **NOAC**<br>**monotherapy**<br>**(N=126)** | **Combination**<br>**therapy**<br>**(N=121)** | **Hazard ratio**<br>**(95% CI)** | **P-value** | **NOAC**<br>**monotherapy**<br>**(N=356)** | **Combination**<br>**therapy**<br>**(N=357)** | **Hazard ratio**<br>**(95% CI)** | **P-value** | **Interaction**<br>**P-value** |
| **Net adverse clinical events\*** | 12 (9.5) | 26 (21.5) | 0.42 (0.21-0.83) | 0.01 | 34 (9.6) | 56 (15.7) | 0.59 (0.39-0.90) | 0.02 | 0.40 |
| Death from any cause | 5 (4.0) | 6 (5.0) | 0.80 (0.24-2.62) |  | 9 (2.5) | 13 (3.6) | 0.69 (0.30-1.62) |  |  |
| Cardiovascular death | 2 (1.6) | 3 (2.5) | 0.64 (0.11-3.83) |  | 5 (1.4) | 8 (2.2) | 0.63 (0.20-1.91) |  |  |
| Myocardial infarction | 0 | 3 (2.5) | NA |  | 4 (1.1) | 3 (0.8) | 1.34 (0.30-5.98) |  |  |
| Stent thrombosis | 0 | 1 (0.8) | NA |  | 2 (0.6) | 1 (0.3) | 2.01 (0.18-22.12) |  |  |
| Stroke | 3 (2.4) | 0 | NA |  | 2 (0.6) | 4 (1.1) | 0.50 (0.09-2.73) |  |  |
| Major bleeding | 2 (1.6) | 13 (10.7) | 0.14 (0.03-0.62) |  | 9 (2.5) | 16 (4.5) | 0.55 (0.24-1.23) |  |  |
| Clinically relevant<br>nonmajor bleeding | 4 (3.2) | 8 (6.6) | 0.4 (0.13-1.45) |  | 10 (2.8) | 26 (7.3) | 0.37 (0.18-0.78) |  |  |
| **Ischemic composites†** | 5 (4.0) | 6 (5.0) | 0.81 (0.25-2.65) | 0.73 | 11 (3.1) | 13 (3.6) | 0.85 (0.38-1.90) | 0.69 | 0.95 |
| **Bleeding composites‡** | 6 (4.8) | 21 (17.4) | 0.25 (0.10-0.63) | <0.01 | 19 (5.3) | 42 (11.8) | 0.44 (0.26-0.76) | <0.01 | 0.31 |

NA denotes not applicable. Abbreviations as in **Table 1**.
\*Net adverse clinical events were defined as a composite of all-cause death, myocardial infarction, stent thrombosis, stroke (ischemic and hemorrhagic), systemic embolism, or major bleeding or clinically relevant nonmajor bleeding. Systemic embolism was not reported in any patients in either group.
†Ischemic composites were defined as a composite of cardiovascular death, myocardial infarction, stent thrombosis, ischemic stroke, or systemic embolism.
‡Bleeding composites were defined as a composite of major or clinically relevant nonmajor bleeding.

Ischemic outcomes were infrequent and did not differ significantly between NOAC monotherapy and combination therapy in either the complex PCI group (4.0% vs 5.0%; HR, 0.81; 95% CI, 0.25–2.65; *P*=0.73) or the noncomplex PCI group (3.1% vs 3.6%; HR, 0.85; 95% CI, 0.38–1.90; *P*=0.69), with no evidence of interaction (*P*_interaction_=0.95). In the complex PCI group, the absolute risk difference for ischemic outcomes was −0.99%.

Bleeding outcomes were consistently lower with NOAC monotherapy than with combination therapy in both the complex PCI group (4.8% vs 17.4%; HR, 0.25; 95% CI, 0.10–0.63; *P*<0.01) and the noncomplex PCI group (5.3% vs 11.8%; HR, 0.44; 95% CI, 0.26–0.76; *P*<0.01), without a significant interaction by PCI complexity (*P*_interaction_=0.31). Among patients with complex PCI, major bleeding occurred less frequently with NOAC monotherapy than with combination therapy (1.6% vs 10.7%; HR, 0.14; 95% CI, 0.03–0.62; *P*<0.001).

### Subgroup analyses by the number and components of complex PCI features

Among the 126 patients in the complex PCI group, 62 (49%) had a single complex feature and 64 (51%) had multiple complex features. NOAC monotherapy was associated with a lower risk of NACE in both subgroups (single feature: adjusted HR, 0.24; 95% CI, 0.07–0.87; *P*=0.03; multiple features: adjusted HR, 0.21; 95% CI, 0.08–0.59; *P*<0.01) (**Figure 4**). Similar patterns were observed for bleeding outcomes, whereas ischemic outcomes did not differ significantly between treatment strategies.

**Figure 4.**
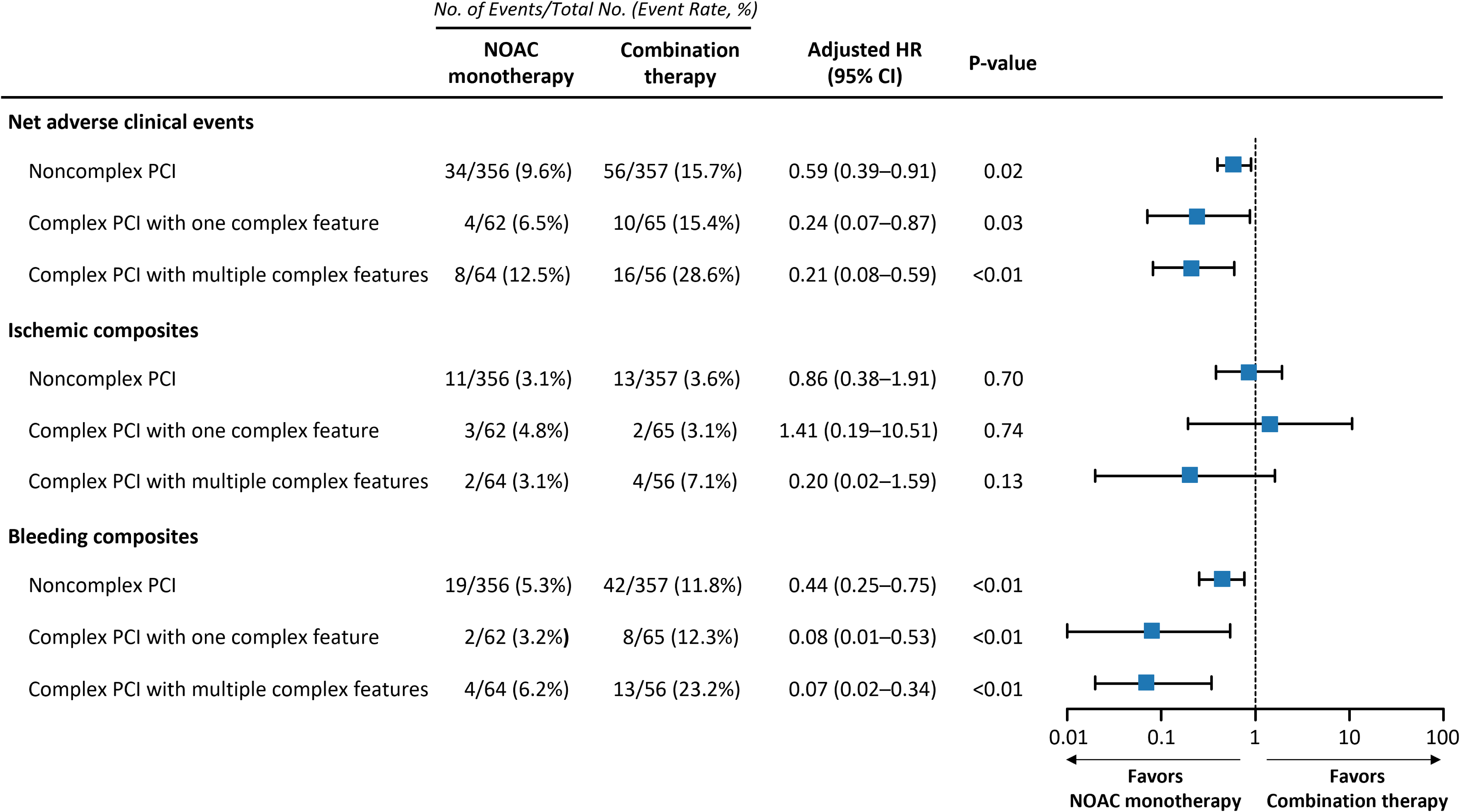
Forest plot according to the number of complex PCI features. Hazard ratios for NOAC monotherapy relative to combination therapy were estimated using Cox proportional hazards models and are presented with 95% confidence intervals. Multivariable models were adjusted for clinically relevant covariates. Abbreviations as in **Figure 1 and 2**.

Analysis according to each individual complexity component are presented in **Supplementary Figure S1**. NOAC monotherapy was associated with a net clinical benefit in patients with ≥3 stents implanted and in those with a total stent length ≥60 mm. For other complexity components, including ≥3 lesions treated, bifurcation PCI with two stents, left main PCI, and chronic total occlusion PCI, NOAC monotherapy showed numerically lower rates of NACE, although these differences did not reach statistical significance, likely owing to limited sample size and low event rates.

## DISCUSSION

In this post-hoc analysis of the ADAPT AF-DES trial, we compared NOAC monotherapy with combination therapy consisting of a NOAC plus clopidogrel in patients with AF stratified by PCI complexity. The main findings of the current study are as follows. First, NOAC monotherapy was associated with a significantly lower risk of net adverse clinical events at 12 months compared with combination therapy in both the complex and noncomplex PCI groups. Second, NOAC monotherapy was not associated with an increased risk of ischemic events, regardless of PCI complexity. Third, among patients with complex PCI, NOAC monotherapy was associated with a marked reduction in bleeding events.

Although current guidelines recommend oral anticoagulant monotherapy in patients with AF and stable CAD beyond 1 year after PCI, uncertainty has remained regarding its applicability in patients undergoing complex PCI, who are traditionally considered to be at heightened ischemic risk.^3,4^ Importantly, prior randomized trials supporting oral anticoagulant monotherapy did not systematically evaluate anatomic PCI complexity. The AFIRE and EPIC-CAD trials enrolled patients with heterogeneous revascularization strategies without explicit lesion-based stratification, while complex PCI was not prespecified or systematically categorized in OAC-ALONE.^1,2,5^ In AQUATIC, although complex PCI was included, patient-related high-risk features were incorporated into eligibility criteria, limiting lesion-specific inference.^13^ Accordingly, the present analysis addresses a clinically relevant evidence gap by evaluating antithrombotic strategies in patients with anatomically complex PCI.

Consistent with the overall trial results, NOAC monotherapy was associated with a significant reduction in bleeding among patients with complex PCI. Although no formal interaction was observed between PCI complexity and antithrombotic strategy, the absolute bleeding reduction was numerically greater in the complex PCI group than in the noncomplex PCI group, largely driven by fewer major bleeding events. These findings are clinically relevant, given that complex PCI has been consistently associated with increased bleeding risk in addition to ischemic risk.^14,15^ Within this context, the bleeding reduction observed with NOAC monotherapy appears particularly meaningful in routine practice, although subgroup comparisons should be interpreted cautiously owing to limited event numbers.

Importantly, NOAC monotherapy was not associated with an excess risk of ischemic events, even among patients with complex PCI. Ischemic event rates were low, and no numerical signal of increased ischemic risk was observed across individual complexity components or among patients with multiple complex features. Although the precision of effect estimates was limited by the small number of events, the absence of a consistent ischemic signal supports the safety profile observed in the parent trial.

This neutral ischemic effect may reflect contemporary PCI practice. Traditionally, prolonged or intensive antithrombotic therapy after complex PCI has been justified by procedural considerations, including extensive stent implantation and treatment of anatomically complex lesions, which were regarded as mediators of stent-related thrombotic risk.^6,16^ However, advances in stent technology—such as thinner struts and biocompatible polymers—together with improvements in PCI techniques and intravascular imaging-guided optimization strategies, have substantially mitigated stent-driven ischemic complications.^17-21^ In contrast to first-generation drug-eluting stents, for which the cumulative incidence of very late stent thrombosis beyond 1 year after PCI was reported to be as high as 1.6% over 5 years, very late stent thrombosis has become rare with second- and third-generation drug-eluting stents.^7,22-25^ Moreover, several studies of antiplatelet strategies have shown no excessive increase in ischemic events, even in the setting of complex PCI.^26,27^ Consistent with these observations, the incidence of stent thrombosis in the complex PCI subgroup of the present analysis was extremely low. Although detailed information on intravascular imaging use was not available in the ADAPT AF-DES trial, the relatively high adoption of imaging-guided PCI in Korea may partly explain the low incidence of stent-related thrombotic events observed in the present study.^28^

In the present study, patients with certain high bleeding risk features, such as a history of recurrent bleeding or baseline anemia, were proactively excluded.^8^ Nevertheless, higher bleeding vulnerability was observed among patients with complex PCI. Because bleeding events related to prolonged antiplatelet therapy are strongly linked to adverse prognosis, including increased mortality,^29^ and AF further amplifies bleeding risk in patients with CAD, a bleeding-focused antithrombotic strategy may be particularly appropriate in this population.^30,31^

### Limitations

Several limitations of the present study should be acknowledged. First, this was a post hoc analysis and was not powered for subgroup comparisons, particularly for individual complexity components; therefore, these findings should be regarded as hypothesis-generating. Second, PCI complexity was not prespecified in the original trial protocol and was defined retrospectively, raising the possibility of residual confounding despite methodological consistency with prior studies.^8,32^ Third, the definition of complex PCI used in this study may differ from those applied in other studies, especially regarding lesion type and complexity thresholds.^10^ Fourth, this analysis predominantly reflects treatment with apixaban and rivaroxaban; therefore, extrapolation to other NOACs should be made with caution. Fifth, the trial population consisted exclusively of East Asian patients, who are known to differ from Western populations in terms of both ischemic and bleeding susceptibility; consequently, the generalizability of our findings to other ethnic groups may be limited.^20,33^ Despite these limitations, in the absence of high-quality randomized evidence specifically addressing antithrombotic strategies after complex PCI in patients with AF, the present analysis provides clinically relevant insights and offers an important basis for the design of future prospective studies.

### Conclusions

In this post-hoc analysis of the randomized ADAPT AF-DES trial, NOAC monotherapy beyond 12 months after PCI was associated with a substantial reduction in bleeding without an apparent increase in ischemic events, regardless of PCI complexity. These findings support a bleeding-focused long-term antithrombotic strategy in patients with AF and stable CAD, including those who have undergone complex PCI, while underscoring the need for prospective validation.

## Non-standard abbreviations and acronyms

ADAPT AF-DES: Appropriate duration of antiplatelet and thrombotic strategy after 12 months in patients with atrial fibrillation treated with drug-eluting stents
AF: atrial fibrillation
CAD: coronary artery disease
NACE: net adverse clinical events
NOAC: non–vitamin K antagonist oral anticoagulant
PCI: percutaneous coronary intervention.

## Acknowledgments

The authors reviewed and edited the manuscript and take full responsibility for its content. Artificial intelligence–based language tools were used to assist with language editing.

## Data availability statement

The data supporting this paper will be shared by the corresponding author upon reasonable request.

